# Comparison of foundation models and transfer learning strategies for diabetic retinopathy classification

**DOI:** 10.64898/2026.04.17.26351092

**Authors:** Livie Yumeng Li, Benjamin Lebiecka-Johansen, Stine Byberg, Vajira Thambawita, Adam Hulman

**Affiliations:** Department of Public Health, Aarhus University, Aarhus, Denmark; Steno Diabetes Center Aarhus, Aarhus University Hospital, Aarhus, Denmark; Clinical Epidemiological Research, Copenhagen University Hospital – Steno Diabetes Center Copenhagen, Herlev, Denmark; Simula Metropolitan Center for Digital Engineering, Oslo, Norway

**Keywords:** Deep learning, Foundation models, Calibration, Diabetic retinopathy, Classification

## Abstract

Diabetic retinopathy (DR) is a leading cause of vision impairment, requiring accurate and scalable diagnostic tools. Foundation models are increasingly applied to clinical imaging, but concerns remain about their calibration. We evaluated DINOv3, RETFound, and VisionFM for DR classification using different transfer learning strategies in BRSET (n = 16,266) and mBRSET (n = 5,164). Models achieved high discrimination in binary classification (normal vs retinopathy) in BRSET (AUROC 0.90-0.98), with DINOv3 achieving the best under full fine-tuning (AUROC 0.98 [95% CI: 0.97-0.99]). External validation on mBRSET showed decreased performance for all models regardless of the fine-tuning strategy (AUROC 0.70–0.85), though fine-tuning improved performance. Foundation models achieved strong discrimination but poor calibration, generally overestimating DR risk. While the generalist model, DINOv3, benefited from deeper fine-tuning, miscalibration remained evident. These findings underscore the need to improve calibration and the comprehensive evaluation of foundation models, which are essential in clinical settings.

**Author summary:** Artificial intelligence is increasingly being used to detect eye diseases such as diabetic retinopathy from retinal images. Recent advances have introduced “foundation models,” which are trained on large datasets and can be adapted to new tasks. We aimed to evaluate how well these models perform in a clinical prediction context, with a focus not only on accuracy but also on how reliably they estimate disease risk.

In this study, we compared different types of foundation models using two independent datasets from Brazil. We found that while these models were generally good at distinguishing between healthy and diseased eyes, their predicted risks were often poorly calibrated. In other words, the estimated probabilities did not consistently reflect the true likelihood of disease.

We also examined whether adapting the models to the target population could improve performance. Although this approach led to improvements, calibration issues remained. However, post-training correction improved the agreement between predicted risks and observed outcomes.

Our findings highlight an important gap between model performance and clinical usefulness. We suggest that improving the reliability of risk estimates is essential before such systems can be safely used in healthcare.

## Introduction

Diabetic retinopathy (DR) is the most common microvascular complication of diabetes mellitus and a leading cause of preventable vision loss among working-age people.^1^ Regular screening with retinal fundus imaging enables early detection and timely treatment, reducing severe outcomes.^2,3^ Images are graded with standardized procedures to detect DR severity and progression.

Foundation models mark a major advance in deep learning, trained on large, non-annotated datasets for a general purpose. In image analysis, foundation models are typically pre-trained on large generic (e.g., ImageNet) or domain-specific datasets and can be fine-tuned for specific tasks.^4^ These models learn general visual representations that can be tailored to clinical applications and used as decision support, which enable transfer across imaging modalities and disease areas.^5,6^ In ophthalmology, models like RETFound and VisionFM have been trained on millions of ophthalmic images to support downstream tasks such as DR classification.^7,8^

As the complexity and size of state-of-the-art models increase, training a model from scratch requires large-scale datasets and substantial computational resources. It has become essential to apply transfer learning, which inherits some of the model’s parameters and layers that are pre-trained on large datasets and adapts them to specific new objectives using various updating strategies.^9,10^ These strategies vary in the extent of model adjustment, from updating only the last fully connected layers (head fine-tuning) to fine-tuning the entire model (full fine-tuning). Depending on the pre-training domain and model architecture, the choice of strategy may also impact performance and generalizability.^11^

Discrimination is the most commonly reported predictive performance metric.^12^ It measures how well a model distinguishes between individuals with and without a specific outcome. Calibration, which assesses how well predicted probabilities match actual outcomes, is particularly important when guiding individual-level decisions.^13,14^ However, calibration is often underreported in deep learning research.^15^ This is particularly concerning as modern deep neural networks tend to exhibit poor calibration despite strong discrimination, highlighting the urgent need to investigate calibration of foundation models.^16–18^ Decision curve analysis, which assesses clinical utility by calculating net benefit across different risk thresholds, is also rarely reported in deep learning studies.^19,20^

Therefore, this study aimed to evaluate and compare foundation models for DR prediction from retinal fundus images using transfer learning in both binary and multiclass settings, with a special focus on metrics beyond discrimination, especially calibration.

## Methods

### Study population and data collection

The primary dataset used in the study is the Brazilian Multilabel Ophthalmological Dataset (BRSET), which was collected from three outpatient ophthalmological centers in São Paulo, Brazil, between 2010 and 2020.^21,22^ This project was approved by the São Paulo Federal University institutional review board (CAAE 33842220.7.0000.5505). The requirement for individual consent was waived. In this dataset, all images were anonymized with all identifiable patient information removed. This dataset comprises 16,266 single 45° retinal fundus images from 8,524 individuals, 16% of whom had diabetes. Images were taken with two retinal cameras, either Canon CR2 or Nikon NF5050. Images were labeled by a retinal specialist according to predefined protocols.^23^ For external evaluation, we utilized the Portable Retina Fundus Photos Dataset (mBRSET), which was collected using a portable, handheld retinal camera, the Phelcom Eyer, from participants attending the Itabuna Diabetes Campaign held in November 2022 in Itabuna, Bahia State, Northeast Brazil.^24,25^ This study was approved by the Instituto de Ensino Superior Presidente Tancredo de Almeida Neves institutional review board (CAAE 64219922.3.0000.9667). All participants provided written informed consent. This dataset included 5,164 fundus images (single 45° images) from 1,291 participants, 97% of whom had diabetes. Images were independently labeled by two ophthalmologists following the same predefined protocols as in BRSET.^23^

In both datasets, image grading followed the International Clinical Diabetic Retinopathy severity scale, which categorizes images into five levels: no retinopathy (Grade 0), mild non-proliferative DR (NPDR, Grade 1), moderate NPDR (Grade 2), severe NPDR (Grade 3), and proliferative DR (Grade 4).^26^ In this study, grades were combined in two ways: (1) binary classification distinguishing normal (Grade 0) from DR (Grades 1-4); and (2) three-class classification distinguishing between normal (Grade 0), NPDR (Grades 1-3), and proliferative DR (Grade 4).

### Foundation models

We compared the performance of a state-of-the-art generalist foundation model (DINOv3) against eye-specific foundation models (RETFound, VisionFM) and their variants. All models were trained with self-supervised learning strategies.

DINOv3 is the most recent version of the DINO models, which stands for distillation with no labels.^27^ It is a generalist foundation model based on the Vision Transformer (ViT), trained via self-distillation on 17 billion natural images.^28^ Variants scale from small to very huge (up to 7 billion parameters) and learn high-quality dense features applicable to classification, segmentation, and other tasks. In this study, ViT-Large-based DINOv3 was used, with approximately 300 million parameters.

RETFound is an ophthalmology-specific foundation model pre-trained on 1.6 million unlabelled retinal images, including retinal fundus images and optical coherence tomography (OCT) scans.^7^ We evaluated the original RETFound model as well as two additional variants: RETFound-DINOv2-SDPP and RETFound-DINOv2-MEH, which are recent variants based on an earlier DINO backbone pre-trained on retinal images from datasets from Shanghai and the Moorfields Eye Hospital.^29,30^ These variants were included to enable comparison between recent eye-specific foundation model developments and the state-of-the-art generalist foundation model.

VisionFM is a multimodal, multitask foundation model pre-trained on 3.4 million ophthalmic images covering various eye diseases, imaging modalities, devices, and patient demographics.^8^ It was included as a representative alternative ophthalmology foundation model to RETFound, enabling comparison across distinct eye-specific pre-training strategies.

Neither BRSET nor mBRSET was included in the training sets of the included foundation models.

### Experimental strategy

Images lacking DR grading in the raw dataset were excluded prior to dataset stratification (complete-case analysis). Fine-tuning datasets were stratified based on patient ID and DR status, divided into training (70%), validation (20% within the training set), and testing (30%) sets, ensuring that splits were mutually exclusive at the patient level. The training set was used for model training, the validation subset guided early stopping and model optimization, and the held-out test set was used solely for final evaluation.

Images were resized to 256×256 pixels to ensure a standard size, then randomly cropped to the models’ required input size of 224×224 pixels per epoch to increase data variability.

We then evaluated model performance in three experiments. In Experiment 1, the pre-trained foundation models were fine-tuned to the BRSET training set for DR classification. We compared performance between head and full fine-tuning strategies. In an exploratory analysis, feature embeddings were extracted from the test set and visualized using uniform manifold approximation and projection (UMAP) to examine differences in representation associated with the fine-tuning strategy. The visualisation was not used for quantitative evaluation.

To evaluate the generalizability of the foundation models fine-tuned to BRSET, in Experiment 2, we performed external evaluation on the independent mBRSET without additional retraining. This external evaluation reflects how differences in diabetes and DR prevalence, and imaging equipment influenced the results.

In Experiment 3, we further investigated whether and how transfer learning can improve generalizability by fine-tuning the models on the mBRSET training set. Each model retained its original fine-tuning strategy from Experiment 1, i.e., models previously head fine-tuned to BRSET were head fine-tuned to mBRSET, and models fully fine-tuned were fully fine-tuned again. Performance was evaluated on the held-out mBRSET test set.

All experiments were conducted in both binary and three-class classification settings. Binary classification results are presented in the main text, with three-class results provided in the Supplementary Materials.

### Predictive performance evaluation

Discrimination was evaluated using the area under the receiver operating characteristic curve (AUROC). Discrimination metrics for three-class classification, including one-vs-rest AUROC and the polytomous discrimination index (PDI), are reported in the Supplementary Materials.

Predicted probability distributions were visualized using histograms. To account for class imbalance and enhance interpretability, counts in each probability bin were normalized by the total number of cases per class. Calibration was assessed with calibration curves (using LOESS smoother),^31^ calibration intercept and slope, and expected calibration error (ECE). Calibration intercept reflects systematic over- or underestimation of risk, and the calibration slope reflects the spread of predicted probabilities. ECE summarizes the mismatch between predicted probabilities and observed outcomes across probability bins and is frequently used in the data science literature.^32^ In addition, calibration was evaluated using equal-mass (decile-based) binning to mitigate binning sensitivity.

To address potential miscalibration, we applied two post-hoc recalibration methods: Platt scaling and temperature (slope-only) scaling.^18,33^ Recalibration parameters were estimated on 1/3 of the test set and subsequently applied to and evaluated in the remaining 2/3.

Differences (Δ) in performance metrics between models and training strategies were estimated using paired bootstrap resampling (n=1,000) of the test set at the image level.

To assess performance at clinically relevant thresholds, sensitivity, specificity, positive predictive value (PPV), and negative predictive value (NPV) were computed for binary classification. Clinical utility was further evaluated using decision curve analysis.^19,20^ Net benefit was calculated across clinically relevant thresholds and compared with default strategies of treating none or all.

### Computational setup

Experiments and analyses were performed using Python (v3.12.5) and R (v4.4.3). A summary of image preprocessing, data augmentation, and model training settings is provided in Supplementary Table 1, in line with reproducibility recommendations for deep learning in medical imaging.^34^ Detailed information on software versions, packages, and code for reproducing the results is provided in our GitHub repository (https://github.com/hulmanlab/brset_mlcp).

## Results

### Study population

In the BRSET, the median (Q1, Q3) age of participants was 61 (47, 71), and female participants constituted 61% of the dataset (Table 1). The prevalence of diabetes was 16% (1,344 out of 8,524 participants), and DR was present in 1,083 of the 16,266 retinal images (7%).

**Table 1.**
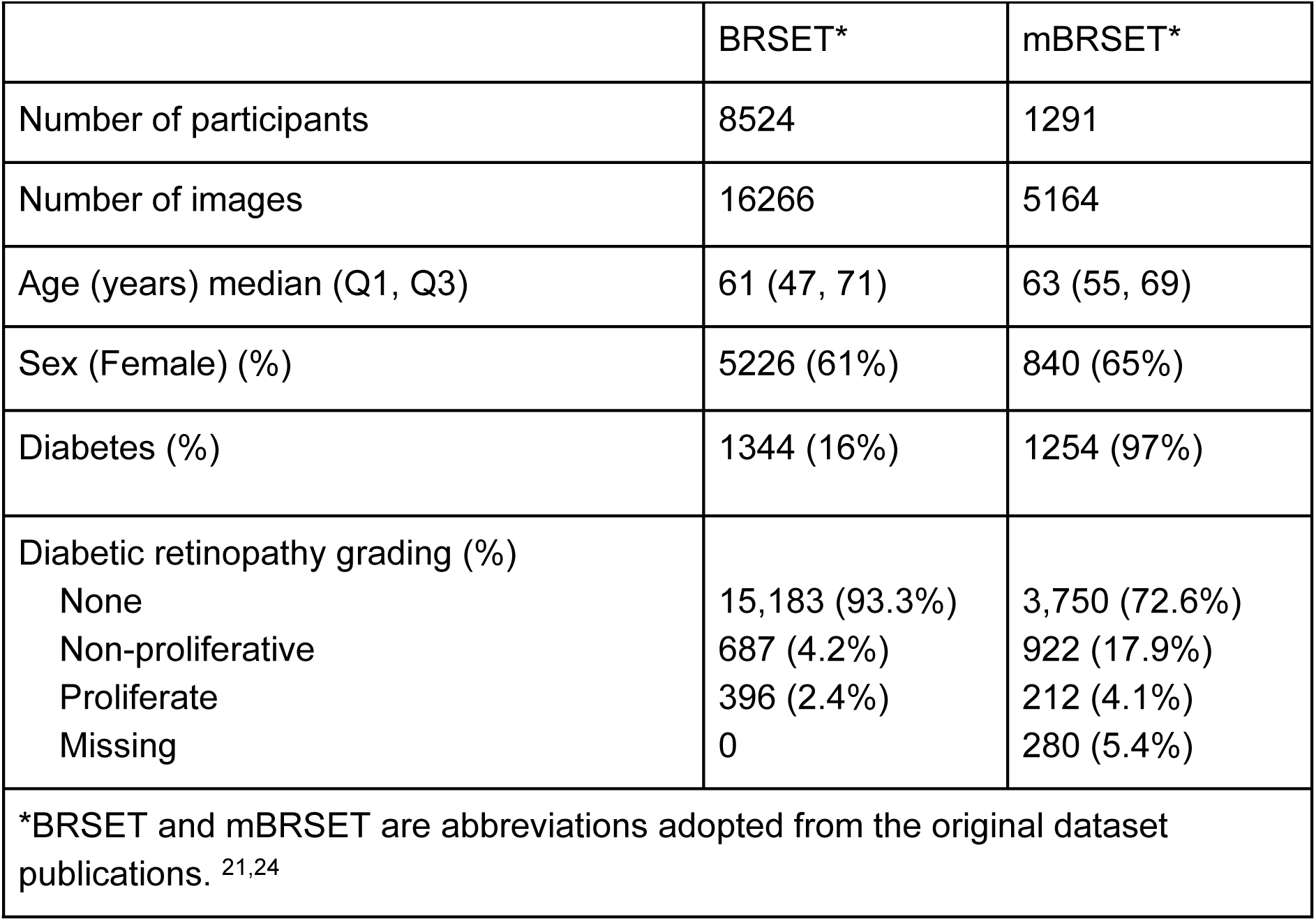
Characteristics of the study population.

In the mBRSET, the median age of participants was 63 (55, 69), and female participants constituted 65% of the dataset. The prevalence of diabetes was 97% (1,254 out of the 1,291 participants). Of the 5,164 retinal images, 280 lacked DR grading and were excluded; among the remaining images, DR was present in 1,134 (23%).

### Discrimination performance

#### Generalist vs eye-specific foundation model

In binary classification, the generalist foundation model DINOv3 achieved a higher AUROC than eye-specific foundation models across all three experimental settings (Figure 1 and Supplementary Table 2). When full fine-tuned on BRSET, DINOv3 achieved an AUROC of 0.98 [0.97, 0.99], compared with 0.95 [0.93, 0.96] for RETFound and 0.90 [0.87, 0.92] for VisionFM, corresponding to differences in AUROC of 0.03 [0.02, 0.05] and 0.09 [0.06, 0.11], respectively. During external validation in mBRSET, all models had lower AUROC by 0.12 to 0.25, but DINOv3 still outperformed RETFound and VisionFM by 0.13 [0.11, 0.15] and 0.11 [0.09, 0.12], respectively. Transfer learning to mBRSET improved model AUROC by 0.04 to 0.10, but DINOv3 continued to outperform the eye-specific models.

**Figure 1.**
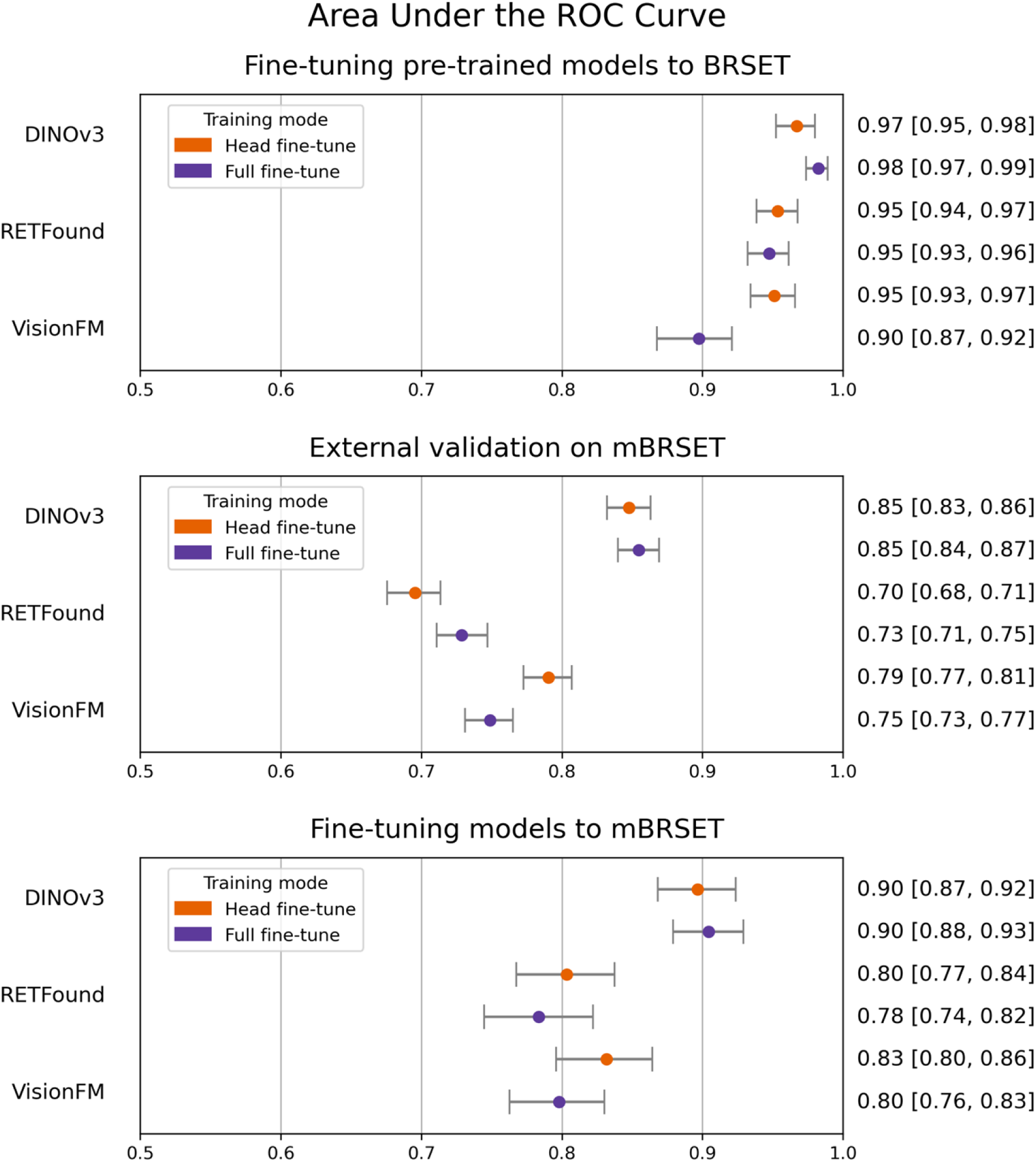
Discrimination performance of generalist and eye-specific foundation models.

Additional comparisons included recent eye-specific RETFound variants, RETFound-DINOv2-SDPP and RETFound-DINOv2-MEH (Supplementary Table 3). DINOv3 achieved a higher AUROC when full fine-tuned on BRSET (Supplementary Table 4). During external validation on mBRSET and after fine-tuning BRSET-trained models to mBRSET, DINOv3 also achieved higher AUROC with differences ranging from 0.01 to 0.07.

#### Head vs. full fine-tuning

Discrimination performance differed between head and full fine-tuning strategies only for the eye-specific foundation models (Supplementary Table 5). For RETFound, AUROC did not differ between fine-tuning strategies when models were trained on BRSET. However, full fine-tuned RETFound achieved a higher AUROC than head fine-tuning during external validation on mBRSET (ΔAUROC: 0.03 [0.02, 0.05]). In contrast, for VisionFM, full fine-tuning consistently achieved lower AUROC than head fine-tuning across all three experiments, with differences ranging from −0.03 to −0.05.

The feature visualization of embeddings extracted from head fine-tuned models showed distinct clustering by fundus camera type (Figure 2). Feature projections from different camera types were less separated after full fine-tuning.

**Figure 2.**
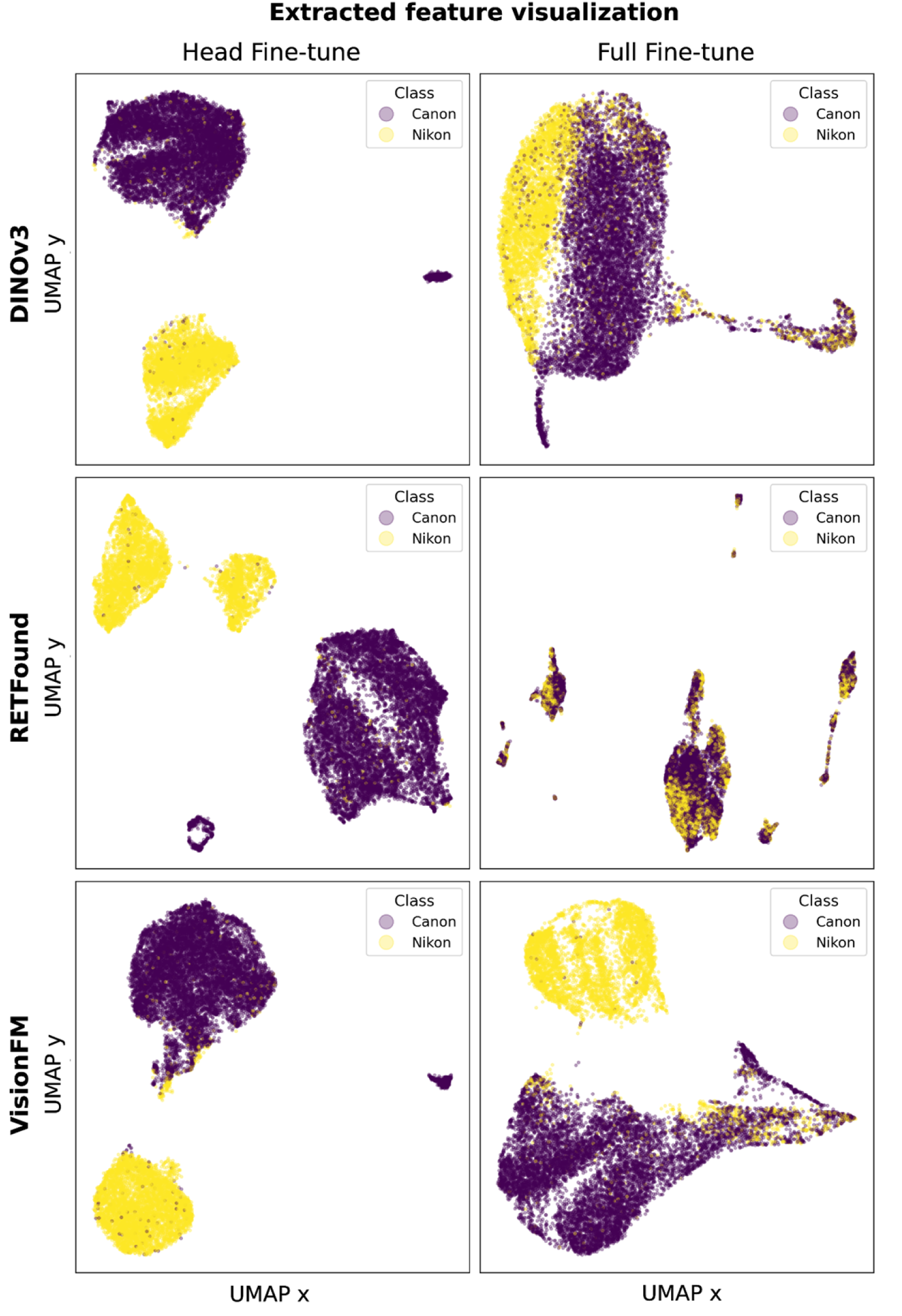
Feature space visualization of head and full fine-tuned foundation models by camera type.

### Calibration performance

Despite high discrimination performance, all models showed substantial miscalibration (Figures 3 and 4, Supplementary Table 6). For DINOv3, predicted probabilities for normal images were concentrated between 0.1 and 0.4, with average predicted risks of 15% and 16% in the lowest decile for head and full fine-tuned models respectively, despite having no observed DR. For RETFound, probabilities assigned to normal images concentrated closer to zero, with average predicted risk of 5% and 12% in the lowest decile for head and full fine-tuned models respectively, but were at least 2% with head fine-tuning and 8% with full fine-tuning for those without DR. VisionFM showed the most pronounced miscalibration for those without DR, with average predicted risk of 32% and 21% in the lowest decile for head and full fine-tuned models, respectively. Calibration curves, calibration intercept and slope indicated marked deviations from ideal calibration across models (Supplementary Table 6). For fine-tuning on BRSET, all intercepts were below 0, ranging from −4.27 to −1.82, indicating overestimation of risk. Among all models and fine-tuning combinations, head fine-tuned RETFound showed the lowest ECE (0.11 [0.10, 0.12]), consistent with the qualitative assessment of the probability distribution plots (Supplementary Table 6).

**Figure 3.**
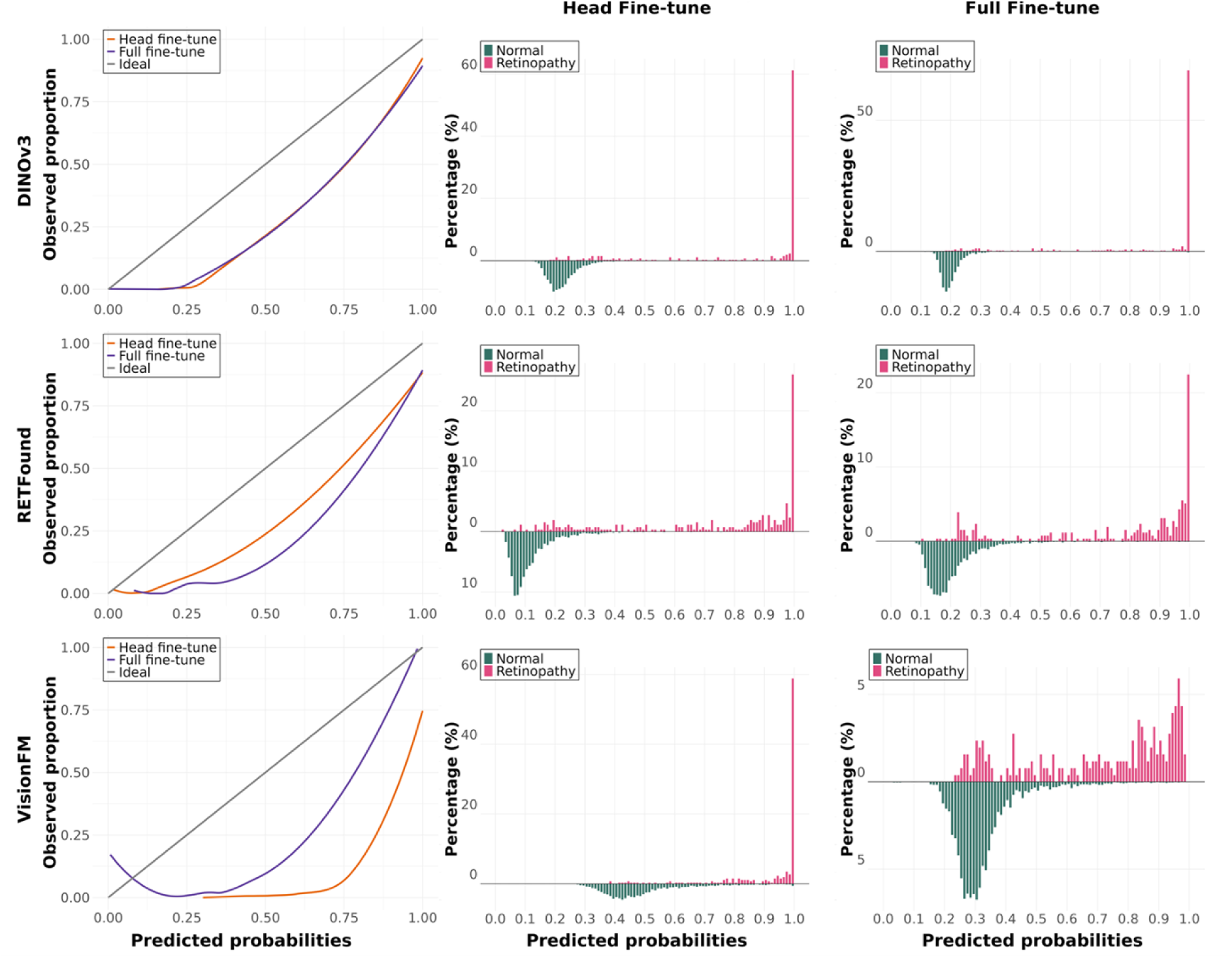
Calibration curves and predicted probability distributions for models fine-tuned on BRSET.

**Figure 4.**
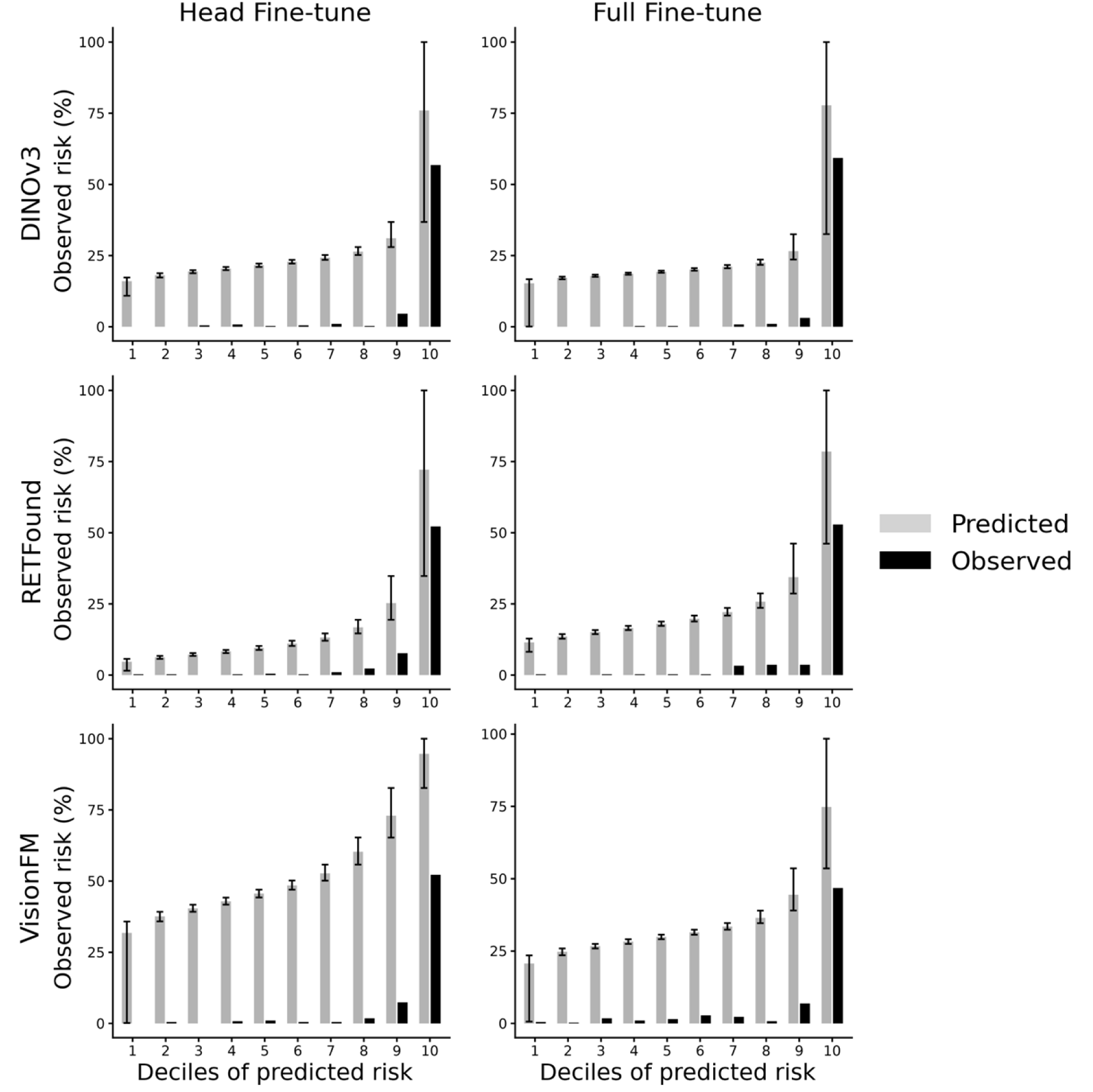
Calibration plot (equal-mass binning) for models fine-tuned on BRSET. Error bars indicate the minimum and maximum observed event rates within each bin.

Calibration performance was next evaluated for external validation on mBRSET and fine-tuning BRSET-trained models to mBRSET (Supplementary Figures 1-4). In both settings, predicted probability distributions and calibration curves were further away from the ideal line compared with fine-tuning on BRSET. External validation showed the largest deviations, while fine-tuning to mBRSET resulted in reduced deviations relative to external validation. During external validation, full fine-tuned RETFound was assessed with the lowest ECE (0.05 [0.05, 0.06], Supplementary Table 6). Despite pronounced deviations in the probability distribution plots, it showed the best alignment among models in the equal-mass binning calibration plot.

### Effect of recalibration

Post-hoc recalibration was evaluated for full fine-tuned DINOv3 across the three experimental settings (Supplementary Figure 5, Supplementary Table 7). Both Platt scaling and temperature scaling improved calibration, with Platt scaling showing greater improvement (calibration intercepts ranging from 0.02 to 0.19, and slopes from 0.92 to 1.08). When fine-tuned to BRSET, recalibration assigned minimal predicted risks across the lowest nine deciles, with correspondingly low or no observed event rates. During external validation and subsequent fine-tuning on BRSET, risk overestimation is reduced by recalibration.

### Clinical utility

When fine-tuned on BRSET, models had very high sensitivity (0.84 to 1.00) and negative predictive value (0.90 to 1.00), but substantially lower specificity (0.00 to 0.87) and positive predictive value (0.06 to 0.52) (Supplementary Table 8). External validation and fine-tuning to mBRSET were associated with higher specificity and positive predictive value compared with fine-tuning on BRSET, while negative predictive value remained high across settings.

Decision curve analysis was performed for full fine-tuned DINOv3 across the three experimental settings before and after recalibration using Platt scaling (Supplementary Figure 6). When fine-tuned on BRSET, the model’s net benefit decreased below 0 from a threshold of 0.07 while overlapping with the ‘treat all’ strategy until 0.15, corresponding to the minimum predicted probability. After recalibration, the model’s net benefit exceeded both reference strategies across a broad range of threshold probabilities of interest. During external validation on mBRSET and after fine-tuning BRSET-trained models to mBRSET, the model’s net benefit also exceeded both reference strategies.

Results from the three-class classification task are summarised in Supplementary Figures 7-9 and Supplementary Tables 9-11.

## Discussion

This study compared the performance of foundation models for predicting DR in binary and multiclass classification settings using different transfer learning strategies. Foundation models achieved strong discrimination but consistently exhibited poor calibration. Transfer learning improved discrimination when adapting models to a new dataset, which differed in population characteristics, imaging equipment, and outcome distribution. The state-of-the-art generalist foundation model, DINOv3, outperformed the eye-specific foundation models.

In this study, all models exhibited miscalibration despite strong discrimination performance. Poor calibration is reflected in overestimating individual risk, causing unnecessary referrals, missed interventions, and increased patient burden and healthcare costs. For example, DINOv3 assigned a 10-40% risk of retinopathy to normal images in BRSET, illustrating the potential for over-referral due to miscalibration. Threshold-specific metrics showed very high sensitivity and NPV, alongside comparatively lower specificity and PPV. This pattern reflected that miscalibrated probabilities exceeded the decision threshold even for images without DR, potentially resulting in unnecessary referrals or follow-ups for healthy patients and increasing the burden on the healthcare system.

Most studies in clinical prediction research report discrimination but not calibration metrics, and these practices are unfortunately not different for DR grading.^21,24^ In clinical practice, predicted probabilities and risk scores are often compared with a certain threshold to guide clinical decisions.^35,36^ Therefore, calibration affects how model outputs translate into referral decisions. This dependence on decision thresholds is further reflected in the decision curve analysis, where net benefit is calculated directly from predicted probabilities at thresholds. When full fine-tuning DINOv3 to BRSET, the net benefit was negative between threshold 7% and 15%, indicating more harm than benefit. This range corresponded to the minimum predicted risk among those without DR. After Platt scaling recalibration, its net benefit exceeded ‘treat all’ and ‘treat none’ strategies across thresholds. The shift in predicted probabilities caused by miscalibration can alter net benefit estimates and affect conclusions regarding clinical utility.

Post-hoc recalibration corrected miscalibration to some extent, with Platt scaling showing the greatest improvement. However, the magnitude of correction differed across recalibration methods and experiments. These findings indicate that although miscalibration can be mitigated post-hoc, calibration behavior remains sensitive to both the recalibration approach and the experimental context. Alternative approaches to improve calibration beyond post-hoc adjustment, including data augmentation and loss function modifications, should also be considered.^37,38^

Classical calibration metrics suggested improved calibration under external validation, similar to findings from a prior methodological study that reported improved ECE under distribution shift.^37^ However, metrics such as calibration intercept and slope are based on fitting parametric models, essentially a line, and can be misleading when predicted probabilities are highly concentrated. This issue became more pronounced in the multiclass setting, where miscalibration occurred between adjacent stages, when calibration estimates for proliferative DR were based on limited case numbers. Probability distribution plots revealed a more complex picture and complemented classical calibration metrics, supporting the recent guidance on the importance of reporting such plots.^15^

Greater attention should be given to clinical prediction models’ calibration, as well as their context of use, such as threshold selection and decision rules, which is consistent with TRIPOD-AI guidance on reporting model usability in practice.^12^ This consideration is particularly relevant for deep learning models, which are known to be susceptible to miscalibration.^37^

External validation is essential to assess a model’s generalizability. A recent study by Xiong et al. externally evaluated RETFound on a dataset from China and highlighted its limited generalizability, underscoring challenges for efficacy and equity.^39^ In Experiment 2, we externally validated a model trained on BRSET, a dataset from the general population collected in outpatient ophthalmological centers, using the mBRSET, which consisted of individuals with diabetes and was collected using a hand-held retinal imaging device at a diabetes campaign. Performance decreased likely due to differences in dataset characteristics and outcome prevalence, which highlights the importance of validating models under conditions that mirror their intended use, particularly when translating models from resourceful clinics to resource-limited settings. Despite the overall performance decline, DINOv3 exhibited the smallest decrease in AUROC under external validation, suggesting greater robustness when the backbone pre-trained on the huge natural image dataset was preserved.

In Experiment 3, we applied an additional transfer learning step by fine-tuning the models to mBRSET. Fine-tuning improved discrimination across all models and training strategies compared with direct external validation in Experiment 2. This pattern highlights that transfer learning to a related target domain can effectively mitigate domain shift and enhance discrimination.

The state-of-the-art generalist foundation model, DINOv3, generally achieved the highest AUROC across experiments and training strategies when compared with the established eye-specific foundation models RETFound and VisionFM. We also compared DINOv3 with the most recent publicly available RETFound variants (RETFound-DINOv2), where their performance was comparable when fine-tuned on BRSET, whereas under external validation and subsequent adaptation to mBRSET, DINOv3 maintained better discrimination.

In foundation models, the backbone network contains knowledge inherited from the pre-training process, while the classification head adapts this knowledge to the specific task. Full fine-tuning updates both components, offering deeper adaptation to the new task, but requiring more computational resources. Across all experiments, we observed little to no difference in AUROC between head and full fine-tuning. Exploratory feature visualization further showed that although backbone representations captured device-related characteristics, fine-tuning aligned the model with the classification task, and device differences were smaller in the feature space. Prior evidence suggests that full fine-tuning consumes more computational resources than full fine-tuning, indicating that the choice of transfer learning strategy should be informed by adaptation needs and resource constraints to translate deep learning models into scalable clinical applications.^40,41^

Our estimates of model discrimination align with previous studies evaluating DINOv2, RETFound, and VisionFM for DR detection on benchmark datasets, despite differences in datasets and experimental settings.^7,8^ Similarly, when compared with studies using the BRSET and mBRSET datasets, our results are broadly consistent with previously reported discrimination metrics.^21,24^ Beyond using only images, a recent foundation model, EyeCLIP, employs multimodal contrastive learning and can incorporate textual information in its predictions, opening new possibilities for multimodal clinical decision support.^42^ Foundation models have also been applied to other clinical data modalities, including dermatology and radiology images,^43,44^ as well as time-series data such as electrocardiography and continuous glucose monitor data.^45–47^ Reporting of model calibration is generally rare across domains, revealing an important gap in the evaluation of foundation models for clinical use and the importance of our findings beyond ophthalmology.

### Strengths and limitations

This study offers methodological insights into using foundation models in the clinical setting. We compared models pre-trained on general vs. eye-specific datasets, using head or full fine-tuning in binary and multiclass classification. We evaluated discrimination, calibration with probability distribution plots, and decision curve analysis to give a comprehensive comparison of performance. External validation using a dataset including almost exclusively people with diabetes enabled a realistic assessment of how transfer learning can adapt models to new, different settings. Notably, we offer novel evidence on the calibration performance of foundation models for DR classification and mitigation strategies to tackle miscalibration. While most studies focus only on simplifying DR severity into binary categories, we included a three-class approach to retain more detail. Although DR severity is inherently ordinal, our evaluation framework did not explicitly incorporate ordinal-aware metrics, which limits our ability to distinguish adjacent from cross-stage misclassifications. However, this approach still does not reflect clinical practice and may reduce sensitivity to early-stage disease. In addition, although exploratory analyses suggest that device-related variation is less prominent after fine-tuning, we cannot determine whether the observed external miscalibration is primarily driven by differences in imaging devices or by differences in outcome prevalence between datasets.

## Conclusion

This study demonstrated the potential and limitations of foundation models for DR classification. While foundation models achieved strong discrimination, poor calibration remained a key limitation. Although good discrimination is valuable for ranking individuals by risk, it is insufficient when absolute risk is required for decision-making on an individual level. Different transfer learning strategies yielded different results depending on the pre-training domain. Applying transfer learning improved discrimination and calibration, highlighting its value in domain adaptation. Future research should focus on evaluating these methods in real-world clinical settings, including calibration of foundation models in ophthalmology and other clinical domains. By evaluating state-of-the-art foundation models using a more comprehensive clinical assessment framework, this study helps bridge the gap between advances in AI model development and clinically oriented model evaluation.

## Data Availability

The datasets used in the study are open access and are available on PhysioNet (BRSET: https://doi.org/10.13026/1pht-2b69, mBRSET: https://doi.org/10.13026/qxpd-1y65). The models and code from the study are available on GitHub (https://github.com/hulmanlab/brset_mlcp).

https://physionet.org/content/mbrset/1.0/

https://physionet.org/content/brazilian-ophthalmological/1.0.1/

## Data and code availability

The datasets used in the study are open access and are available on PhysioNet (BRSET: https://doi.org/10.13026/1pht-2b69, mBRSET: https://doi.org/10.13026/qxpd-1y65).^22,25,48^ The models and code from the study are available on GitHub (https://github.com/hulmanlab/brset_mlcp).^49^

## Acknowledgment

We would like to thank GenomeDK (https://genome.au.dk/) and Aarhus University for providing computational resources and support that contributed to these research results.

## Author contributions

LYL, SB, VT, and AH contributed to the conceptualization and design of the study. LYL acquired and prepared the data, set up the model pipeline, and implemented the experiments. LYL and BLJ contributed to technical validation. LYL, BLJ, and AH contributed to evaluation methodologies. LYL and AH interpreted the results. AH, SB, and VT supervised the study. LYL drafted the initial manuscript. All authors critically revised the manuscript and approved the final version.

